# Validation of NG-TEST^®^ *Acineto*-5^®^ for the Detection of OXA Carbapenemases, NDM, and VIM in the *Acinetobacter calcoaceticus-baumannii* Complex and beyond

**DOI:** 10.64898/2026.10.19.26365660

**Authors:** Saoussen Oueslati, Alphée Piard, Arnaud Chalin, Laura Francius, Anaïs Vogel, Margaux Le Quellec, Emeric Dupre, Christian Moguet, Stéphanie Simon, Hervé Volland, Katy Jeannot, Thierry Naas

**Affiliations:** Team ‘Resist’, UMR1184 ‘Infectious disease models and innovative therapies (IDMIT)’ INSERM, Université Paris-Saclay, CEA, Health, Faculty of Medicine, Le Kremlin-Bicêtre, France; Department of Bacteriology-Hygiene, Bicêtre Hospital, APHP Paris-Saclay, Le Kremlin-Bicêtre, France; French National Reference Center for Antibiotic Resistance: Pseudomonas and Acinetobacter, Besançon, France; Department of Bacteriology CHU Besançon Franche-Comté, Besançon, France; Research and development department, NG Biotech, Guipry, France; Université Paris-Saclay, CEA, INRAE, Département Médicaments et Technologies pour la Santé, 91191, Gif-sur-Yvette, France; SEPSIS Comprehensive Center – IHU SEPSIS, France; French National Reference Center for Antibiotic Resistance: Carbapenem-Resistant Enterobacterales, Le Kremlin-Bicêtre, France

**Keywords:** CHDL, OXA-carbapenemase, Acinetobacter, detection, multiplex LFIA

## Abstract

Carbapenem resistance in *Acinetobacter* spp. is frequently mediated by acquired carbapenemases, whose rapid detection can inform antimicrobial therapy and infection-control decisions. NG-TEST^®^ *Acineto*-5^®^ is a lateral flow immunoassay designed to detect major acquired carbapenemases in *Acinetobacter* spp. We evaluated its performance using 433 whole-genome-sequenced isolates, including single producers of OXA-23-like (n = 126), OXA-24-like (n = 14), OXA-58-like (n = 8), OXA-143/253/542 (n = 10), VIM-like (n = 9), and NDM-like enzymes (n = 25), as well as isolates co-producing two (n = 20) or three (n = 2) carbapenemases. The collection was predominantly composed of *Acinetobacter baumannii* (n = 365) and *Acinetobacter pittii* (n = 33), with 13 additional species represented by 1 to 7 isolates each. The assay detected all targeted carbapenemases, including all tested variants and combinations, yielding 100% sensitivity. OXA-24-like, OXA-143-like, and the distantly related OXA-542 were detected by the same antibody pair, demonstrating cross-reactivity within this group. No false-positive results occurred among isolates lacking targeted carbapenemases, resulting in 100% specificity. Weak positive signals were observed in four of five *A. radioresistens* isolates carrying intrinsic *bla*_OXA-23-like_ genes, consistent with low-level expression. Overall, NG-TEST® *Acineto*-5^®^ enables rapid detection of major acquired carbapenemases in *Acinetobacter* spp., supporting infection control, epidemiological surveillance and therapeutic decision-making, as carbapenemase-producing isolates are often multidrug resistant. Detection of VIM and NDM may be particularly valuable because these enzymes are not inhibited by durlobactam and have been associated with increased cefiderocol MICs, potentially limiting treatment options.

**IMPORTANCE:** Carbapenem-resistant *Acinetobacter* spp. are important causes of healthcare-associated infections and are frequently multidrug resistant, leaving few therapeutic options. The diversity of acquired carbapenem-hydrolyzing class D β-lactamases (CHDLs) and metallo-β-lactamases (MBLs) complicates their rapid phenotypic detection. We evaluated the NG-TEST^®^ *Acineto*-5^®^ lateral flow immunoassay using a large, whole-genome-sequenced collection representing diverse carbapenemase variants and clinically relevant *Acinetobacter* spp. The assay reliably detected all targeted carbapenemases, including OXA-23-, OXA-24-, OXA-58-, OXA-143/253/542-, VIM-, and NDM-type enzymes, as well as isolates producing multiple carbapenemases. These findings support the use of NG-TEST^®^ *Acineto*-5^®^ as a rapid diagnostic tool for detecting *Acinetobacter*-specific carbapenemases, facilitating infection-control measures and antimicrobial therapy. This is particularly relevant with the use of novel β-lactamase inhibitors such as durlobactam in combination with sulbactam. Durlobactam inhibits class D OXA-type carbapenemases but not MBLs such as NDM and VIM; therefore, rapid identification of the underlying carbapenemase mechanism may help inform appropriate therapeutic decisions.

## Introduction

*Acinetobacter* spp. are Gram-negative, non-fermenting bacteria increasingly associated with healthcare-associated infections, with *A. baumannii* being the predominant clinically relevant species (1, 2). Other members of the *A. calcoaceticus–baumannii* (ACB) complex, including *A. pittii*, *A. nosocomialis*, and *A. seifertii*, are also important opportunistic pathogens, particularly in critically ill and immunocompromised patients (1). Their phenotypic similarity can complicate species identification, while their capacity to acquire antimicrobial resistance further increases their clinical importance.

*A. baumannii* is particularly well adapted to the hospital environment, where persistence on surfaces and patient colonization facilitate transmission (3). It causes pneumonia, bloodstream, wound, urinary tract, and other invasive infections, particularly in intensive care and among patients with prolonged hospitalization or invasive devices (2, 4). Bloodstream infections and ventilator-associated pneumonia require rapid and accurate diagnosis to enable timely therapy and reduce mortality (5-8). The increasing prevalence of multidrug-resistant, extensively drug-resistant, and carbapenem-resistant *A. baumannii* (CRAB) has substantially limited treatment options and increased morbidity, mortality, and healthcare burden (1, 2, 4). Accordingly, CRAB is classified by the World Health Organization as a critical-priority pathogen (9).

Carbapenem resistance in *A. baumannii* is mediated by multiple mechanisms, with carbapenem-hydrolyzing β-lactamases being particularly important. Carbapenem-hydrolyzing class D β-lactamases (CHDLs, or OXA-carbapenemases) predominate, particularly OXA-23-like enzymes, followed by OXA-24-like and OXA-58-like enzymes (1, 10, 11). Metallo-β-lactamases (MBLs), particularly NDM and VIM, are also increasingly reported. The intrinsic *bla*_OXA-51_-like gene may contribute to resistance when overexpressed through insertion sequences such as IS*Aba1* (1, 12).

The emergence of CRAB has increased the importance of novel therapies, including cefiderocol and sulbactam-based treatment such as sulbactam-durlobactam. Durlobactam inhibits class A, C, and D β-lactamases, including CHDLs, but does not inhibit MBLs such as NDM and VIM (13). Similarly, cefiderocol generally shows greater *in vitro* activity against CHDL-producing *A. baumannii* than against MBL-producing isolates, although resistance can occur in both groups (14). Thus, identification of the underlying carbapenemase mechanism has increasing clinical relevance beyond simply establishing carbapenem resistance.

Rapid detection of carbapenem resistance and its mechanism can support therapeutic decision-making, antimicrobial stewardship, infection control, and surveillance (15). Conventional antimicrobial susceptibility testing (AST) remains essential but does not reliably identify the underlying mechanism (9). Phenotypic carbapenemase assays may show variable performance, particularly for CHDLs (15, 16), whereas molecular methods such as PCR and LAMP provide rapid and specific gene detection but require specialized equipment and may not reflect gene expression or the resistance phenotype (9). Only a few commercial molecular assays include *Acinetobacter*-associated carbapenemases, and molecular panels have predefined target coverage and may therefore miss clinically relevant *Acinetobacter* carbapenemases (17, 18). Whole-genome sequencing provides comprehensive characterization but remains less suitable for routine diagnostics because of its cost, infrastructure requirements, and bioinformatic complexity (19).

Lateral flow immunochromatographic assays (LFIAs) provide rapid carbapenemase detection with minimal equipment and short turnaround times (20). Although the widely used NG-TEST^®^ CARBA-5^®^ assay (NG-Biotech, Guipry, France) detects the major carbapenemases in Enterobacterales and *Pseudomonas aeruginosa*, it has limited utility for *Acinetobacter* spp. because it does not target the CHDLs that predominate in this genus (21, 22). Consequently, *Acinetobacter*-specific LFIAs targeting these CHDLs, as well as NDM, have been developed. Early assays included detection of OXA-23, OXA-24, OXA-58, and NDM (23), while others specifically targeted OXA-23-producers, the main CHDL, directly from positive blood cultures (24). More recently, RESIST ACINETO (Coris BioConcept, Gembloux, Belgium) has been commercialized for simultaneous detection of OXA-23, OXA-24/58, and NDM from bacterial colonies and positive blood cultures (25, 26). However, the reported occurrence of VIM- and OXA-143-producing *A. baumannii* supports the inclusion of these carbapenemases in molecular screening strategies to ensure a broader detection spectrum (13, 27-31).

Therefore, rapid, sensitive, specific, and broadly applicable methods for carbapenemase detection in *Acinetobacter* spp. remain needed. Here, we evaluated the NG-TEST^®^ *Acineto*-5^®^ (NG-Biotech) for the detection of OXA-23, OXA-24, OXA-58, OXA-143, NDM, and VIM carbapenemases in isolates belonging to the *A. calcoaceticus–baumannii* (ACB) complex and beyond.

## MATERIALS AND METHODS

### Ethics statement

All experiments were performed in compliance with French and European regulations on the care of laboratory animals (European Community [EC] Directive 86/609, French Law 2001-486, 6th June 2001) and with the agreements of the ethics committee of the Commissariat à l’Energie Atomique (CEtEA “Comité d’Ethique en Expérimentation Animale” n° 44) no. 12-026 and 15-055 delivered to S. Simon by the French Veterinary Services and CEA agreement D-91-272-106 from the Veterinary Inspection Department of Essonne (France).

### β-Lactamase purification

The *bla*_OXA-24,_ *bla*_OXA-58_, and *bla*_OXA-143_ gene fragments corresponding to the mature β-lactamases were cloned into the expression vector pET41b (+) (Novagen, VWR International, Fontenay-sous-Bois, France) as previously described (32). OXA-24, OXA-58 and OXA-143 were expressed and purified with the hexa-histidine tag on its C-terminus in one step pseudo-affinity chromatography using a NTA-Nickel column followed by a gel filtration step (GE Healthcare, Les Ulis, France) as previously described (32).

### Monoclonal antibodies and assay development

Biozzi mice (10 weeks old) were immunized with the recombinant OXA-24, OXA-58 and OXA-143 β-lactamase as previously described (33). Twenty monoclonal antibodies (mAbs) of each immunization were tested and the pairs of antibodies showing the best limit of detection (LOD) on OXA-24, OXA-58 and OXA-143 -expressing bacteria were selected and subsequently produced on a large scale. These anti OXA-24, OXA-58 and OXA-143 antibodies together with those targeting OXA-23 (24), NDM, and VIM (33) that have previously been described and generated by OXA-23, NDM-1 and VIM-2 recombinant proteins immunization of mice, respectively, were used for the development of this multiplex assay as previously described (33). As anti-OXA-24 and anti-OXA-143 antibodies showed cross-reactivity, detection of OXA-24 and OXA-143 was combined in a single test line.

### Bacterial strains

Four hundred and thirty-three whole genome-sequenced *Acinetobacter* sp. isolates derived from human clinical samples were used to evaluate the multiplex NG-TEST^®^ *Acineto*-5^®^ LFIA. Isolates collected from specimen from patients with either suspected infection or with suspected colonization by *Acinetobacter* spp are listed in Table 1.

**Table 1:** Tested *Acinetobacter* spp and targeted carbapenemases.

|  | Number of isolates tested | WGS results |  | Targeted carbapenemases identified by WGS |  |  |  |  |  |  |
| --- | --- | --- | --- | --- | --- | --- | --- | --- | --- | --- |
|  |  | Positive<sup>a</sup> | Negative<sup>b</sup> | OXA-23-like | OXA-24-like | OXA-58-like | OXA-143-like | OXA-542-like | NDM-like | VIM-like |
| *A. calcoaceticus/baumannii* complex (ACB) |  |  |  |  |  |  |  |  |  |  |
| *Acinetobacter baumannii* | 365 | 186 | 179 | 135 | 18 | 7 | 2 |  | 38 | 7 |
| *Acinetobacter calcoaceticus* | 2 | 0 | 2 |  |  |  |  |  |  |  |
| *Acinetobacter pittii* | 33 | 9 | 24 |  |  | 1 | 4 | 3 | 2 |  |
| *Acinetobacter lactucae* | 4 | 1 | 3 |  |  |  | 1 |  |  |  |
| *Acinetobacter nosocomialis* | 6 | 2 | 4 |  |  | 2 |  |  |  |  |
| *Acinetobacter seifertii* | 1 | 1 | 0 | 1 |  | 1 |  |  |  |  |
| Total tested ACB | 411 | 199 | 212 | 136 | 18 | 11 | 7 | 3 | 40 | 7 |
| non ACB complex |  |  |  |  |  |  |  |  |  |  |
| *Acinetobacter ursingii* | 7 | 4 | 3 |  |  |  |  |  | 1 | 3 |
| *Acinetobacter bereziniae* | 1 | 1 | 0 |  |  | 1 |  |  |  |  |
| *Acinetobacter haemolyticus* | 1 | 1 | 0 |  |  | 1 |  |  |  |  |
| *Acinetobacter johnsonii* | 1 | 1 | 0 |  |  |  |  |  | 1 |  |
| *Acinetobacter junii* | 3 | 0 | 3 |  |  |  |  |  |  |  |
| *Acinetobacter lwoffii* | 1 | 0 | 1 |  |  |  |  |  |  |  |
| *Acinetobacter variabilis* | 2 | 2 | 0 | 1 |  | 1 |  |  |  |  |
| *Acinetobacter geminorum* | 1 | 1 | 0 |  |  |  | 1 |  |  |  |
| *Acinetobacter radioresistens* | 5 | 5 | 0 | 5 |  |  |  |  |  |  |
| Total tested non-ACB | 22 | 15 | 7 | 6 | 0 | 3 | 1 | 0 | 2 | 3 |
| Total tested *Acinetobacter* spp | 433 | 214<sup>c</sup> | 219 | 142 | 18 | 14 | 8 | 3 | 42 | 10 |

<sup>a</sup>: Number of isolates positive for at least one targeted carbapenemase; <sup>b</sup>: Number of isolates negative for any of the targeted carbapenemase. <sup>c</sup>: Among the 214 isolates positive for at least one targeted carbapenemase, 192 carried a single carbapenemase, 20 carried two targeted carbapenemases, and 2 carried three targeted carbapenemases.

This collection included 411 *Acinetobacter calcoaceticus/baumannii* (ACB) complex isolates with *A. baumannii* (n = 365), *A. calcoaceticus* (n = 2), *A. pittii* (n = 33), *A. lactucae* (n = 4), 1 *A nosocomialis* (n = 6)*, and 1 A. seifertii* (n = 1). Non ACB (n = 22) were also tested, with *A. ursingii* (n=7). *A. bereziniae* (n = 1), *A. haemolyticus* (n = 1), *A. johnsonii* (n = 1), *A. junii* (n = 3), *A. lwoffii* (n = 1), *A. variabilis* (n = 2)*, A. geminorum* (n = 1)*, and A. radioresistens* (n = 5) (Table 1). Among the 214 isolates positive by WGS for one of the targeted carbapenemase, 192 produced a single, and 22 multiple carbapenemases (20 with two and two with three targeted carbapenemases) (Table 1), thus, in total 237 targeted carbapenemase variants were evaluated. Two hundred and nineteen didn’t produce any of the targeted carbapenemases and were used as negative controls. Cross-reactivity of NG-TEST^®^ *Acineto*-5^®^ with various non-targeted oxacillinases found in non-*A. baumannii* and non-ACBs isolates was also tested (Table 1).

The distribution of the tested carbapenemases was consistent with their reported global distribution (4), with OXA-23-like enzymes being the most prevalent (n = 142; 59.9%), followed by NDM-like (n = 42; 17.7%), OXA-24 variants (n = 18; 7.6%), OXA-58 variants (n = 14; 5.9%), VIM variants (n = 10; 4.2%), OXA-143 variants (n = 8; 3.4%), and OXA-542 variants (n = 3; 1.3%), which correspond to OXA-143-related novel acquired OXA-carbapenemase. These data are detailed in Table 1.

### Limit of detection (LOD) using recombinant proteins and bacterial extracts

Serial dilution of the recombinant proteins used for immunization (50,000, to 0 pg/mL) were performed with extraction buffer. Similarly, an OXA-23, -24, -58, -143, VIM-4 and NDM-1 -producing *A. baumannii* isolates (Table 2) were resuspended in water to reach 0.5 Mac Farland (c.a. 10^8^ CFU/mL), and subsequently serially diluted in water (bioMérieux, Marcy l’Etoile, France) (10^7^, 10^6^, 10^5^, 10^4^, 10^3^ CFU/mL). One ml of each dilution was centrifuged for 10 minutes at 10 000 g in a microcentrifuge, and the pellet was resuspended in 150 µL extraction buffer. Consecutively, 100 μL of serially diluted proteins and of bacterial extracts were dispensed onto the NG-TEST^®^ Acineto-5 cassette and results were read by eye after 15 minutes. Hundred microliters of the 10^3^-dilution were plated on Uri4 media (BioRad, Marnes-La-Coquette, France), and colonies were counted the next day to determine the precise bacterial count in each suspension.

**Table 2:** Limit of detection of NG-TEST^®^ *Acineto*-5^®^.

| Carbapenemase | Purified proteins | Bacterial isolates |  |  |  |  |  |
| --- | --- | --- | --- | --- | --- | --- | --- |
|  |  | Strain ID<sup>a</sup> | CFU<br>(band intensity<sup>b</sup>) |  |  |  |  |
| OXA-23 | 3.37 ng/mL | O49H5 | 3.2 10<sup>7</sup><br>(6) | **3.2 10<sup>6</sup>** <sup>c</sup><br>(5) | 3.2 10<sup>5</sup><br>/ | / | / |
| OXA-24 | 0.34 ng/mL | O15F10 | 2.3 10<sup>7</sup><br>(10) | 2.3 10<sup>6</sup><br>(9) | 2.3 10<sup>5</sup><br>(8) | **2.3 10<sup>4</sup>**<br>(4) | 2.3 10<sup>3</sup><br>/ |
| OXA-58 | 0.49 ng/mL | O23E5 | 5.2 10<sup>7</sup><br>(9) | 5.2 10<sup>6</sup><br>(8) | 5.2 10<sup>5</sup><br>(7) | **5.2 10<sup>4</sup>**<br>(4) | 5.2 10<sup>3</sup><br>/ |
| OXA-143 | 0.94 ng/mL | O35B8 | 8.9 10<sup>7</sup><br>(9) | 8.9 10<sup>6</sup><br>(9) | **8.9 10<sup>5</sup>**<br>(8) | 8.9 10<sup>4</sup><br>/ | / |
| NDM-1 | 1.23 ng/mL | O57G3 | 6.7 10<sup>7</sup><br>(7) | **6.7 10<sup>6</sup>**<br>(6) | 6.7 10<sup>5</sup><br>/ | / | / |
| VIM-4 | 0.73 ng/mL | O115B5 | 7.2 10<sup>7</sup><br>(8) | 7.2 10<sup>6</sup><br>(6) | **7.2 10<sup>5</sup>**<br>(4) | 7.2 10<sup>4</sup><br>/ | / |

### Study design

The study was conducted at the University Hospital of Bicêtre, Le Kremlin-Bicêtre, France, and at the French National Reference Center for antibiotic resistance in *Pseudomonas aeruginosa* and *Acinetobacter spp*, located at the University Hospital of Besançon, France.

The goal of this study was to evaluate the diagnostic performance of NG-TEST^®^ *Acineto*-5^®^ rapid diagnostic test (NG-Biotech), for detecting OXA-23-, OXA-24-, OXA-58-, OXA-143-, VIM- and NDM-like carbapenemases from clinical *Acinetobacter* spp isolates. The sensitivity and specificity will be assessed retrospectively by comparing LFIA results with whole genome sequencing (Illumina, Les Ulis, France), used as reference method, using colonies grown on blood agar plates (BioMérieux) from strains isolated from specimen from patients who were either infected or colonized and conserved by the investigator’s laboratories.

Three isolated colonies were touched using a one µL loop and resuspended in 5 drops (*circa* 150 µL) of the extraction buffer and vortexed for a few seconds. Subsequently, 100µL of this extract were dispensed on the LFIA cassettes and the migration was followed for 15 minutes. The results were read by eye by monitoring for the appearance of a red band on the test lines, along with a band corresponding to the control line (Figure 1). Results were read and interpreted blindly by two operators and classified as either positive or negative. The test line intensity was then scored according to the intensity scale ranging from 1 to 10, using the “Gold color card” provided by NG-Biotech (Figure 1). For A*. radioresistens* testing was also performed using a one µl loop full of bacteria, and with a five µl loop full of bacteria. In addition, for the five *A. radioresistens* isolates imipenem susceptibility testing using E-tests (BioMérieux) and imipenem hydrolysis determined using CarbAcineto NP test, were determined as previously described (16, 34).

**Figure 1:**
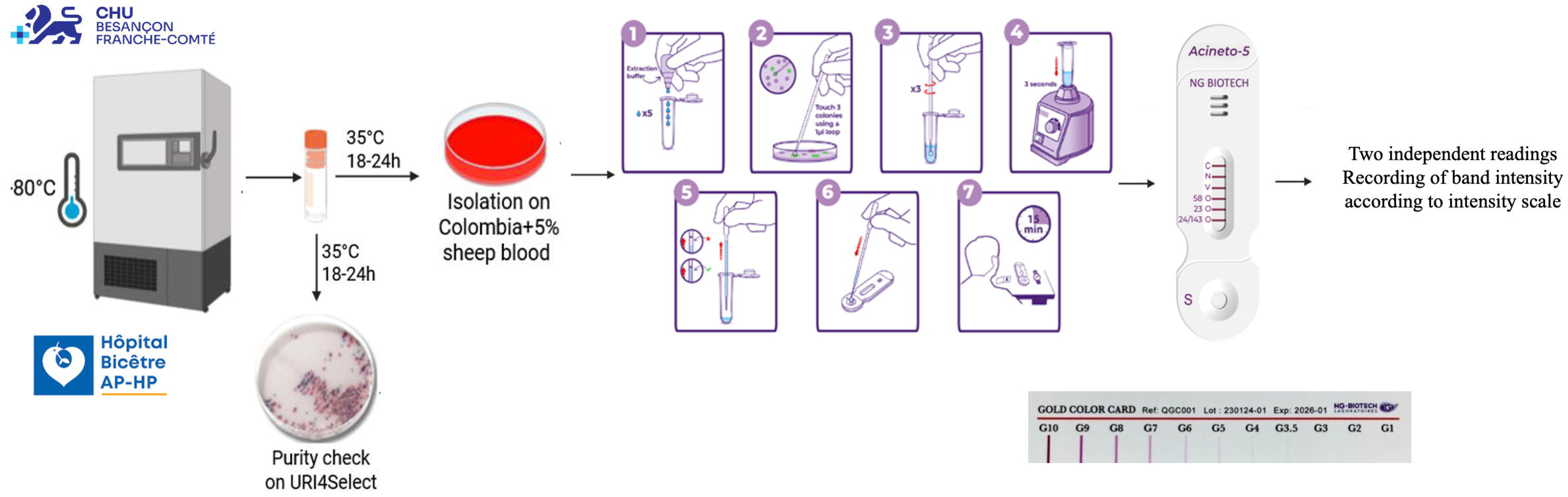
Workflow for the isolation and detection of carbapenemases in *Acinetobacter spp.* clinical isolates. Frozen isolates were recovered by incubation at 35°C for 18–24 h, followed by isolation on Columbia agar supplemented with 5% sheep blood and incubation under the same conditions. Purity was assessed on URI4Select medium. Carbapenemase production was subsequently evaluated using the NG-TEST^®^ *Acineto*-5^®^ assay according to the manufacturer’s instructions, including bacterial extraction, colony suspension, homogenization, sample application, and interpretation after 15 minutes. The assay detects the five targeted carbapenemases represented on the test strip.

### Statistical analysis

The 95% Confidence Intervals (95% CI) were calculated using the online MedCalc statistical software (https://www.medcalc.org/calc/diagnostic_test.php).

## Results

### Distribution of *Acinetobacter* species across the dataset

The *Acinetobacter* spp. isolates evaluated with NG-TEST^®^ *Acineto*-5^®^ (Table 1) originated from diverse clinical specimen, with rectal swabs being the most frequent source (31.5%), followed by urine (20.3%), respiratory specimens (17.4%), blood cultures (16.4%), and from various sources (14.4%).

A heterogeneous distribution of sequence types (STs) was observed according to carbapenemase production. ST2 predominated among carbapenemase-positive isolates (n=81, 37.8 %), while ST2 (n = 24, 10.9 %) and ST1 (n = 9, 4.1 %) were among the most frequent STs in the non-carbapenemase-producing group (Figure 2A). Several STs were shared between groups, whereas others were exclusive to either group, particularly among non-carbapenemase-producing isolates. Overall, the collection showed considerable genetic diversity, with carbapenemase production occurring across multiple ST backgrounds.

**Figure 2.**
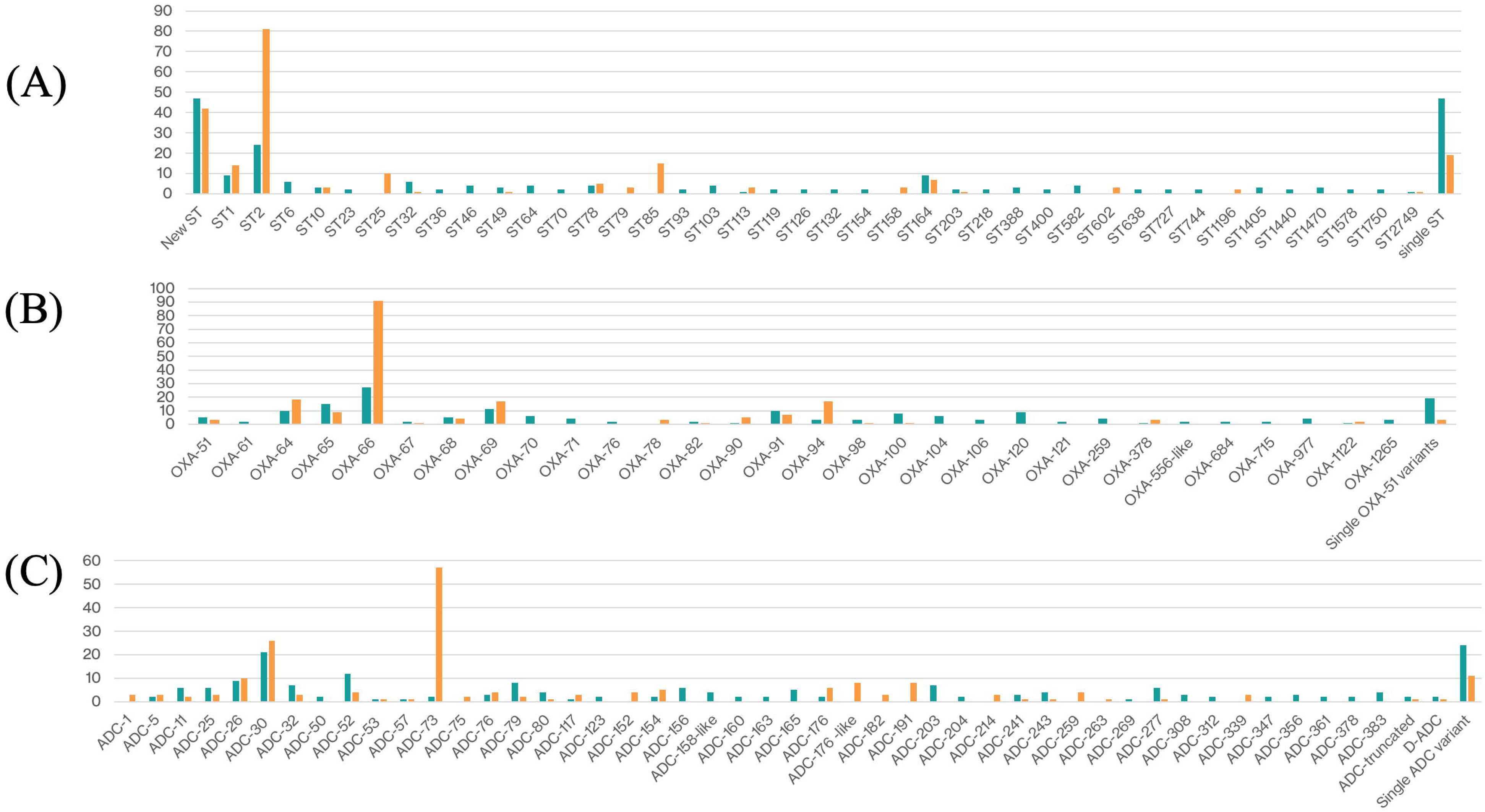
*Acinetobacter* spp. clinical isolates. Distribution of sequence types (STs) (A), of OXA-51 variants (B) and ADC variants (C). “Single ST”, “Single OXA-51 variants” or “Single ADC variants” refer to sequence types, OXA-51 variants and ADC variants detected in only one isolate within the respective category. Green bars represent isolates lacking any of the targeted carbapenemases, whereas those in orange represent isolates producing at least one of the targeted carbapenemases.

In line with the ST diversity observed among the isolates, naturally occurring OXA-51-like and ADC enzymes also displayed substantial diversity (Figure 2B, C). OXA-66 was the most frequent OXA-51-like variant, particularly among carbapenemase-producing isolates followed by OXA-64, OXA-69, OXA-65, and OXA-89, while several additional variants occurred at lower frequencies, including variants found exclusively among non-carbapenemase-producing isolates, such as OXA-120. ADC-30 was detected in both groups, whereas ADC-73 was more frequently associated with carbapenemase-producing isolates; most other variants were identified in only one subgroup. Overall, these findings highlight the broad diversity of naturally occurring ADC enzymes and the underlying β-lactamase heterogeneity among the isolates evaluated.

### LFIA limit of detection (LOD) evaluation

The LOD of NG-TEST^®^ *Acineto*-5^®^ varied according to the carbapenemase tested. Using purified proteins, detection limits ranged from 0.34 to 3.37 ng/mL, with the lowest LOD observed for OXA-24 and the highest for OXA-23 (Table 2). Using *A. baumannii* isolates producing individual carbapenemases, the lowest detectable bacterial concentration ranged from 2.3 × 10⁴ to 6.7 × 10⁶ CFU. The assay showed the highest analytical sensitivity with the OXA-24- and OXA-58-producing isolates, whereas higher bacterial concentrations were required for detection of OXA-23- and NDM-producing isolates. These findings indicate that the analytical detection threshold of NG-TEST^®^ *Acineto*-5^®^ differs according to the carbapenemase enzyme and the bacterial isolate tested.

### Evaluation of multiplex LFIAs with isolates grown on Colombia + 5% sheep blood agar

A total of 433 *Acinetobacter* spp. isolates were evaluated, including 411 (94.9%) belonging to the ACB complex and 365 (84.3%) identified as *A. baumannii* (Table 1). WGS identified at least one targeted carbapenemase in 214 isolates (49.4%), whereas 219 (50.6%) were negative for all targeted carbapenemases. OXA-23-like enzymes were most frequent (n=142), followed by NDM-like (n=42), OXA-24-like (n=18), OXA-58-like (n=14), VIM-like (n=10), OXA-143-like (n=8), and OXA-542-like (n=3) (Tables 1, 3). Among carbapenemase-positive isolates, 192 (89.7%) carried a single targeted carbapenemase, 20 (9.3%) carried two, and 2 (0.9%) carried three (Table 4).

**Table 3:**
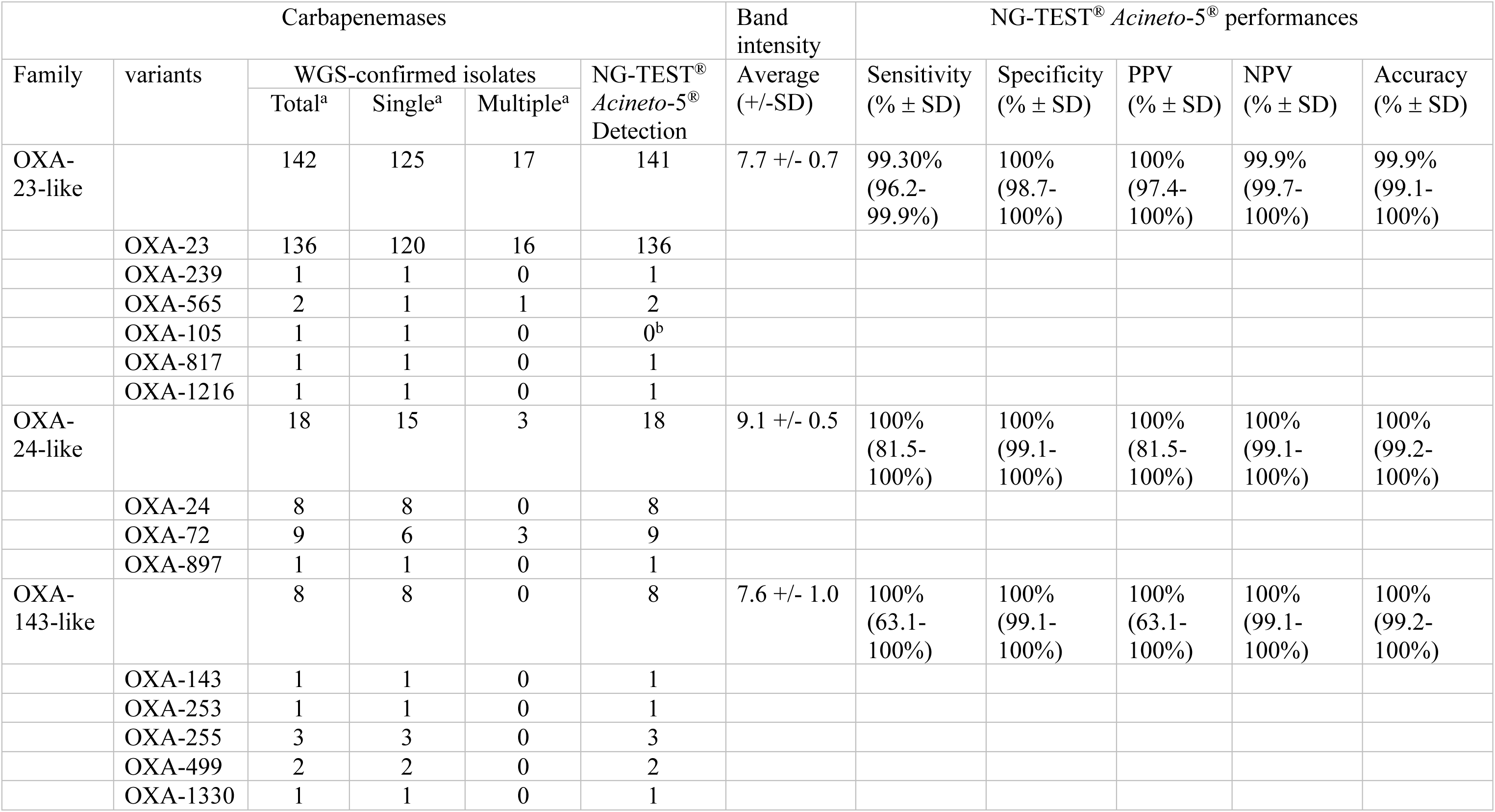
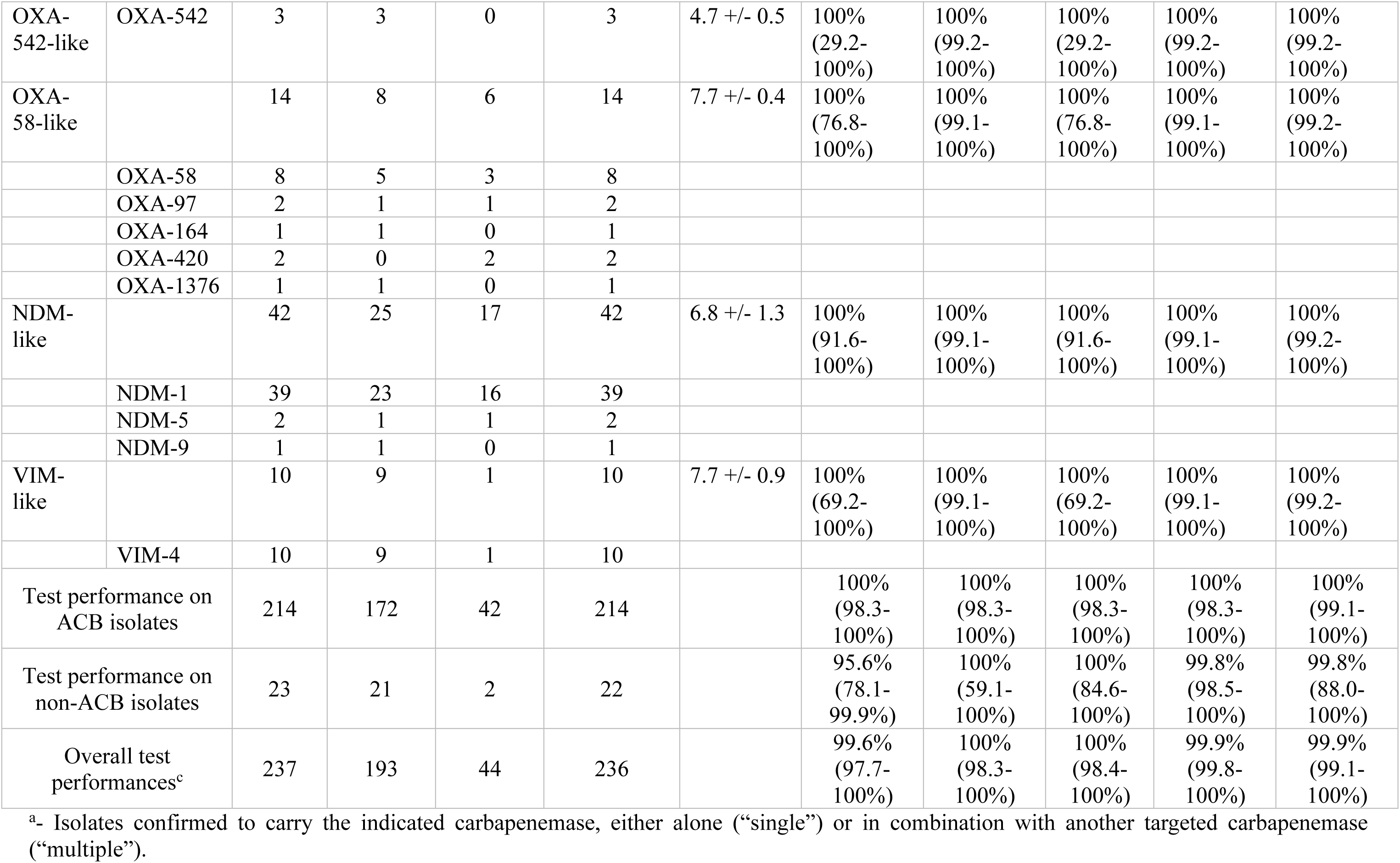
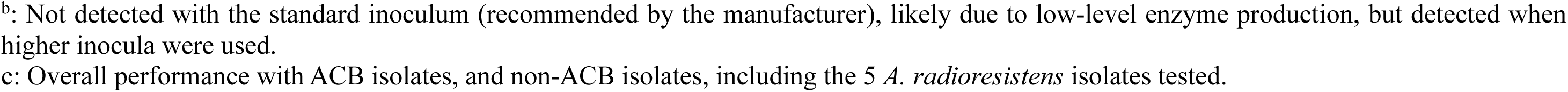
Biological performances of NG-TEST® Acineto 5.

**Table 4:** NG-TEST^®^ *Acineto*-5^®^ results on double and triple carbapenemase producing *Acinetobacter* spp.

| **Double or triple producers** | **Number of isolates** | **NG-TEST® *Acineto-5*® results** |
| --- | --- | --- |
| OXA-23 + OXA-72 (24-like) | 2 | 2 |
| OXA-23 + OXA-97 (58-like) | 1 | 1 |
| OXA-23 + OXA-420 (58-like) | 1 | 1 |
| OXA-23 + NDM-1 | 11 | 11 |
| OXA-565 (23-like) +NDM-1 | 1 | 1 |
| OXA-72 (24-like) + NDM-1 | 1 | 1 |
| NDM-1 + VIM-4 | 1 | 1 |
| OXA-58 + NDM-1 | 2 | 2 |
| OXA-23 + OXA-58 + NDM-5 | 1 | 1 |
| OXA-23 + OXA-420 (58-like) +<br>NDM-1 | 1 | 1 |
| Total | 22 | 22 |

NG-TEST® *Acineto*-5^®^ detected 236 of 237 targeted carbapenemase targets in the 214 WGS-positive isolates, yielding a sensitivity of 99.6% (95% CI, 97.7–100%). Specificity was 100% (95% CI, 98.3–100%), with a PPV of 100% (95% CI, 98.4–100%), NPV of 99.9% (95% CI, 99.8–100%), and overall accuracy of 99.0% (95% CI, 99.1–100%) (Table 3). Targeted carbapenemases were detected across *A. baumannii*, non-*A. baumannii* ACB species (*A. pittii*, *A. lactucae*, *A. nosocomialis*, and *A. seifertii*), and non-ACB species, including *A. ursingii, A. bereziniae, A. haemolyticus, A. johnsonii, A. variabilis, A. geminorum,* and *A. radioresistens* (Table 3).

Most targeted carbapenemases produced band intensities >7 relative to the “Gold color card” reference scale, facilitating interpretation (Figure 1, 3A, 3B; Tables 3). Exceptions included OXA-23 in *A. radioresistens* and OXA-542, which consistently produced weaker signals (intensity 5). OXA-24-like, OXA-143-like, OXA-542-like, OXA-58-like, NDM-like, and VIM-like carbapenemases were detected with 100% sensitivity and specificity. Among OXA-23-like producers, 141/142 were detected (99.3%; 95% CI, 96.2–99.9%). The missed isolate was a carbapenem-susceptible *A. radioresistens* carrying the chromosomally encoded OXA-105. When restricted to ACB complex isolates, both sensitivity and specificity were 100% (Table 3).

**Figure 3:**
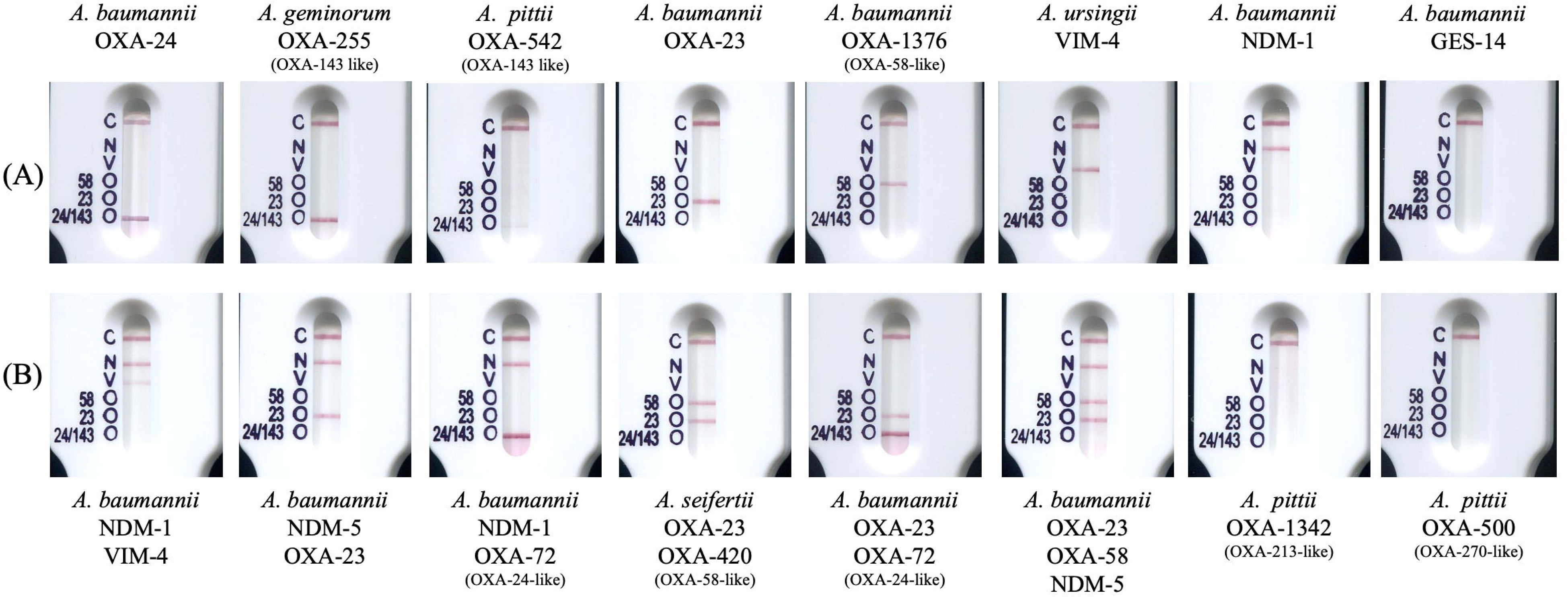
NG-TEST^®^ *Acineto*-5^®^ results for (A) single-carbapenemase producers and (B) multiple-carbapenemase producers. *A. baumannii* GES-14, *A. pittii* OXA-1342 (OXA-213-like), and A*. pittii* OXA-500 (OXA-270-like) illustrate negative results.

Among non-ACB isolates, including the five *A. radioresistens*, sensitivity was 95.6% (95% CI, 78.1–99.9%), while specificity was 100% (95% CI, 59.1–100%), PPV 100% (95% CI, 84.6–100%), NPV 99.8% (95% CI, 98.5–100%), and accuracy 99.8% (95% CI, 88.0–100%).

Only a subset of known variants in the Beta-Lactamase Data Base (http://www.bldb.eu; (35)) was represented: 6/52 OXA-23-like, 3/15 OXA-24-like, 5/9 OXA-58-like, 5/14 OXA-143-like, 3/99 NDM-like, and 1/98 VIM-like variants (Figure 4A–D).

**Figure 4.**
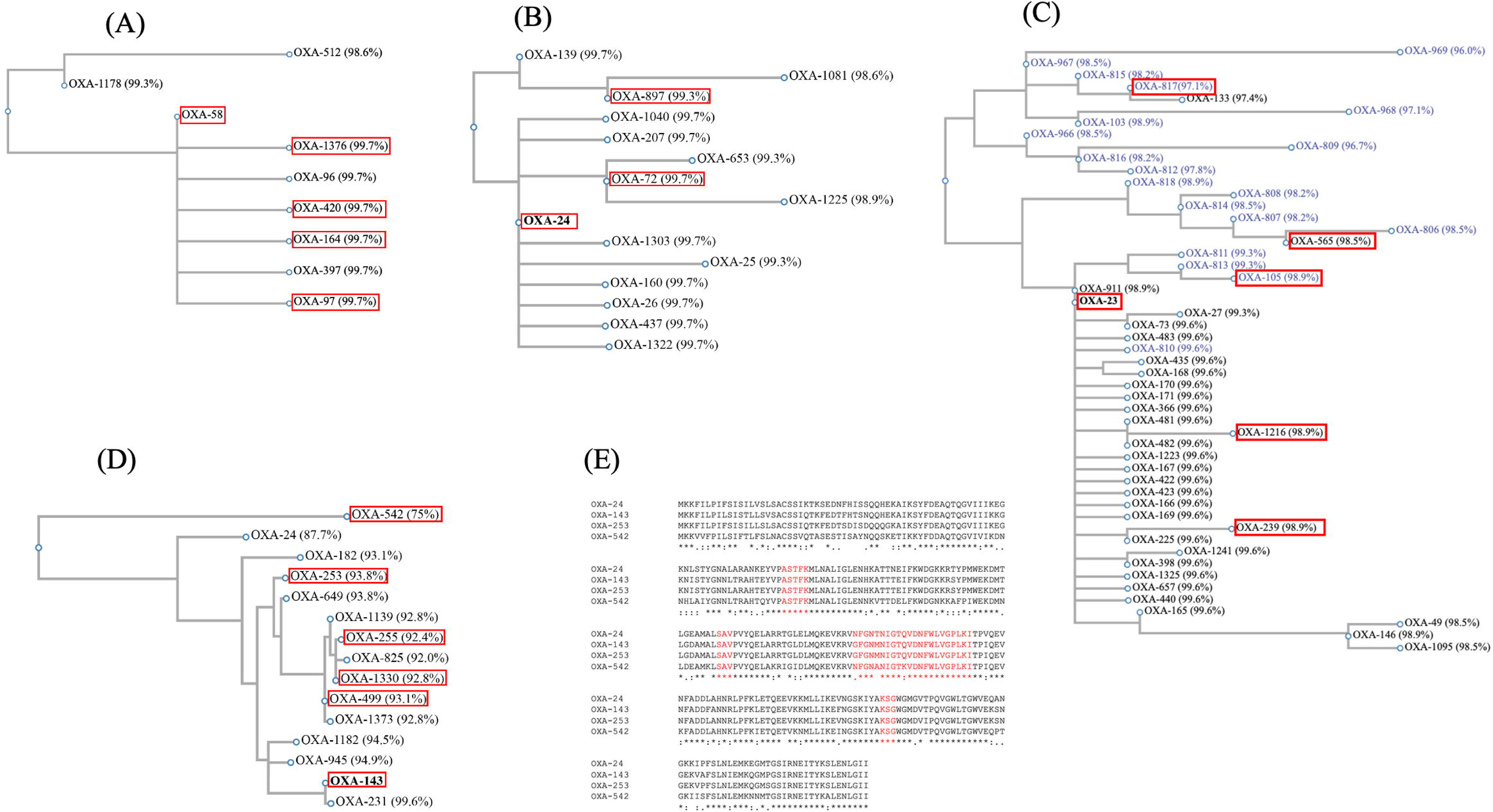
Phylogenetic trees of (A) OXA-58-like, (B) OXA-40-like, (C) OXA-23-like, and (D) OXA-40 and OXA-143-like family members. Red boxes indicate variants tested in this study. (E) Amino acid sequence alignment of OXA-40 with OXA-143, OXA-253, and OXA-542. Conserved residues are indicated by “*”, while conservative amino acid substitutions are indicated by “:” and semi-conservative substitutions by “.” Conserved motifs of class D β-lactamases (45) are indicated in red.

### Detection of OXA-24, OXA-143 and OXA-542

As expected, OXA-24 and OXA-143 variants cross reacted. Indeed OXA-143 shares approximately 88% amino acid identity with OXA-24, which may explain its cross-reactivity with the OXA-24-specific antibodies. They showed equivalent detection of OXA-24 and OXA-143, consistent with the shared antibody recognition of these enzymes (Figure 3A). The assay demonstrated an even broader variant coverage, as OXA-542 that shares approximately 75% amino acid identity with OXA-24 was also detected. OXA-24, OXA-143 and OXA-542 retain substantial stretches of conserved amino acids that may account for its recognition by the OXA-24 antibodies (Figure 4E). This cross-reactivity was associated with a lower band intensity for OXA-542 as compared with the other targeted carbapenemases (Table 3, Figure 3A).

### Evaluation of multiplex LFIAs with multiple carbapenemase producers

A total of 22 isolates (10.3%) identified as multiple carbapenemase producers by WGS, showed concordant detection of the expected carbapenemases by LFIA, corresponding to a 100% detection rate (22/22) (Table 4, Figure 3B). The most frequent co-production profile was OXA-23 + NDM-1, detected in 11 isolates. Other combinations were less frequent, including OXA-23 + OXA-72 and OXA-58 + NDM-1, each identified in two isolates. The remaining profiles, including OXA-23 + OXA-97, OXA-23 + OXA-420, OXA-565 + NDM-1, OXA-72 + NDM-1, NDM-1 + VIM-4, and the two triple-carbapenemase combinations, were each detected in a single isolate. Overall, these findings demonstrate that NG-TEST^®^ *Acineto*-5^®^ LFIA successfully detected the different patterns of carbapenemase co-production among the tested isolates.

### Evaluation of multiplex LFIAs with *A. radioresisens*

Among the five clinical *A. radioresistens* isolates tested, whole-genome sequencing (WGS) identified OXA-23 in three isolates (strains 1, 3, and 5), whereas strains 2 and 4 carried the OXA-23 variants OXA-105 and OXA-817, respectively. Following the manufacturer’s instruction, e.g. touching three colonies, NG-TEST^®^ *Acineto*-5^®^ detected OXA-23 in four isolates out of five with weak band intensities ranging from 3.5 to 5 (Table 5). In contrast, no signal was observed for the OXA-105-producing isolate. Imipenem MICs were in the susceptibility range for all five isolates ranging from 0.25 to 0.75 µg/mL, and imipenem-hydrolytic activity measured using CarbAcineto NP test was negative for all five isolates (Table 5). Notably, increasing the bacterial inoculum (1 µL loop full of bacteria) still resulted in only detection of 4/5 OXA-23-variants, but with slightly stronger band intensities. In contrast using a 5 µL loop full of bacteria allowed detection of all five *A. radioresistens* isolates as OXA-23-producers. The low imipenem MICs and the absence of detectable imipenem-hydrolytic activity support limited OXA-23 expression in these *A. radioresistens* isolates.

**Table 5:** Detection of OXA-23 in *A. radioresistens* by NG-TEST^®^ *Acineto*-5^®^.

| *A. radioresistens* | Isolate | IMP MIC<br>(µg/ml) | CarbAcineto<br>NP test | WGS<br>result | NG-TEST® *Acineto-5*®<br>3 colonies<sup>b</sup> |  | NG-TEST® *Acineto-5*®<br>one µl loop full<sup>c</sup> |  | NG-TEST® *Acineto-5*®<br>5 µl loop full<sup>d</sup> |  |
| --- | --- | --- | --- | --- | --- | --- | --- | --- | --- | --- |
|  |  |  |  |  | Results | Band<br>Intensity | Results | Band<br>Intensity | Results | Band<br>Intensity |
| Strain 1 | O35H1 | 0.5 | Neg<sup>a</sup> | OXA-23 | OXA-23 | 5 | OXA-23 | 6 | OXA-23 | 8 |
| Strain 2 | O27B1 | 0.75 | Neg | OXA-817 | OXA-23 | 5 | OXA-23 | 7 | OXA-23 | 10 |
| Strain 3 | O27E4 | 0.25 | Neg | OXA-23 | OXA-23 | 4 | OXA-23 | 6 | OXA-23 | 8 |
| Strain 4 | O29C4 | 0.25 | Neg | OXA-105 | NEG | - | NEG | - | OXA-23 | 7 |
| Strain 5 | O30G4 | 0.25 | Neg | OXA-23 | OXA-23 | 3.5 | OXA-23 | 5 | OXA-23 | 8 |

<sup>a</sup>: Neg: No hydrolysis detected
<sup>b</sup>: Test results obtained by touching 3 colonies as recommended by the manufacturer (NG-Biotech) and respective band intensities
<sup>c</sup>: Test results obtained using an 1 µL loop full of bacteria and respective band intensities
<sup>d</sup>: Test results obtained using an 5 µL loop full of bacteria and respective band intensities

When *A. radioresistens* isolates were excluded from the analysis, all 232 isolates carrying a targeted acquired carbapenemase were correctly detected, resulting in 100% sensitivity, specificity, positive predictive value, negative predictive value, and overall accuracy (Table 3).

### Evaluation of multiplex NG-TEST^®^ *Acineto*-5^®^ LFIAs with non *A. baumannii* isolates

The cross-reactivity assessment included a broad panel of *Acinetobacter* species carrying diverse naturally occurring and acquired OXA-type carbapenemases, as well as other carbapenemases such as IMP-1 and SIM-1 (Table 6). Despite this genetic diversity, NG-TEST^®^ *Acineto*-5**^®^** showed no cross-reactivity, with negative results for all non-target variants tested. These findings demonstrate high analytical specificity, as the naturally occurring oxacillinases OXA-211-like, OXA-214, OXA-270-like, and OXA-213-like did not interfere with the detection of the targeted carbapenemases (Table 6), despite previous reports of cross-reactivity with the RESIST ACINETO test (Coris BioConcept), particularly for OXA-213 of A*. pittii* (36). Similarly, the recently described acquired carbapenemases OXA-679 and OXA-235, which were not targeted by the assay, were not detected (37, 38).

**Table 6:** NG-TEST^®^ *Acineto*-5^®^ results on *Acinetobacter* spp. clinical isolates producing various naturally occurring oxacillinases.

|  | OXA-134-like |  | OXA-213-like | OXA-270-like | OXA-211-like | OXA-211-like | OXA-229-like | OXA-679-like | WGS confirmed | NG-TEST® *Acineto-5*® result |
| --- | --- | --- | --- | --- | --- | --- | --- | --- | --- | --- |
|  | natural | acquired | natural | natural | natural | natural | natural | acquired | Carbas |  |
| *A. lwoffii* | OXA-283 |  |  |  |  |  |  |  |  | / |
| *A. baumannii* |  | OXA-235 |  |  |  |  |  |  |  | / |
| *A. pittii* |  |  | OXA-417 |  |  |  |  |  |  | / |
| *A. pittii* |  |  | OXA-1266 |  |  |  |  |  | OXA-24 | OXA-24/143 |
| *A. pittii* |  |  | OXA-1331 |  |  |  |  |  | OXA-24 | OXA-24/143 |
| *A. pittii* |  |  | OXA-1342 |  |  |  |  |  |  | / |
| *A. calcoaceticus* |  |  | OXA-844 |  |  |  |  | OXA-946 |  | / |
| *A. calcoaceticus* |  |  | OXA-1050 |  |  |  |  | OXA-1051 |  | / |
| *A. pittii* |  |  |  | OXA-272 |  |  |  |  | SIM-1 | / |
| *A. pittii* |  |  |  | OXA-273 |  |  |  |  | IMP-14<br>OXA-58<br>NDM-1 | OXA-58<br>NDM |
| *A. pittii* |  |  |  | OXA-421 |  |  |  |  |  | / |
| *A. pittii* |  |  |  | OXA-500 |  |  |  |  | IMP-1 | / |
| *A. pittii* |  |  |  | OXA-503 |  |  |  |  | OXA-24 | OXA-24/143 |
| *A. pittii* |  |  |  | OXA-506 |  |  |  |  |  | / |
| *A. pittii* |  |  |  | OXA-564 |  |  |  |  | OXA-24 | OXA-24/143 |
| *A. pittii* |  |  |  | OXA-820 |  |  |  |  |  | / |
| *A. pittii* |  |  |  | OXA-832 |  |  |  | OXA-948 |  | / |
| *A. pittii* |  |  |  | OXA-840 |  |  |  |  | OXA-24 | OXA-24/143 |
| *A. pittii* |  |  |  | OXA-842 |  |  |  |  |  | / |
| *A. pittii* |  |  |  | OXA-1113 |  |  |  | OXA-948 |  | / |
| *A. lactucae* |  |  |  | OXA-678 |  |  |  | OXA-679 |  | / |
| *A. lactucae* |  |  |  | OXA-841 |  |  |  | OXA-949 |  | / |
| *A. lactucae* |  |  |  | OXA-843 |  |  |  | OXA-947 |  | / |
| *A. lactucae* |  |  | OXA-1137 |  |  |  |  |  | OXA-24 | OXA-24/143 |
| *A. johnsonii* |  | OXA-235 |  |  | OXA-333 |  |  |  | NDM-1 | NDM |
| *A. haemolyticus* |  |  |  |  |  | OXA-214 |  |  | OXA-58 | OXA-58 |
| *A. bereziniae* |  |  |  |  |  |  | OXA-301 |  | OXA-58 | OXA-58 |
| *A. geminorum* |  |  |  | OXA-832 |  |  |  |  | OXA-24 | OXA-24/143 |

## Discussion

The present study evaluated the performance of NG-TEST^®^ *Acineto*-5^®^ for rapid detection of major acquired carbapenemases in *Acinetobacter* spp. The assay demonstrated excellent performance, with 99.6% sensitivity and 100% specificity among isolates carrying targeted carbapenemases. Performance was maintained across *A. baumannii*, non-*A. baumannii* ACB complex members, and non-ACB species, which is particularly relevant given the increasing recognition of the diversity of *Acinetobacter* species and carbapenemase determinants encountered in clinical settings (4). Importantly, all 22 isolates producing two or three carbapenemases were correctly detected, including the most frequent OXA-23 + NDM-1 combination. The ability to detect multiple enzymes is particularly relevant in *Acinetobacter*, where co-production of carbapenemases from different families may complicate interpretation of resistance and treatment (39, 40).

The principal advantages of LFIA are its short turnaround time, minimal equipment requirements, and ease of implementation without specialized personnel (20). Compared with molecular methods or WGS, LFIA can provide targeted resistance-mechanism information while comprehensive susceptibility or genomic characterization is pending (20, 21). Rapid carbapenemase detection may therefore support earlier review of antimicrobial therapy and interpretation of microbiological results (41, 42). However, the assay does not replace antimicrobial susceptibility testing, as carbapenemase detection alone does not define the complete susceptibility profile.

Rapid mechanism detection may also support antimicrobial stewardship by facilitating earlier treatment reassessment and avoiding inappropriate reliance on carbapenems. This is particularly relevant with newer agents such as sulbactam-durlobactam, for which durlobactam does not inhibit MBLs, and cefiderocol, which generally shows greater *in vitro* activity against CHDL-producing *A. baumannii* than against MBL producers such as NDM, although resistance may occur in both groups (14). Thus, carbapenemase detection should remain complementary to susceptibility testing.

NG-TEST^®^ *Acineto*-5^®^ may also contribute to infection prevention and control by enabling earlier recognition of carbapenemase-producing *Acinetobacter* spp, which are associated with healthcare transmission and outbreaks (4). Its 100% specificity and absence of cross-reactivity with a broad range of non-target CHDLs further support its potential utility in this setting. Rapid detection could facilitate earlier implementation of appropriate infection-control measures while awaiting more comprehensive characterization.

The genetic diversity observed in the study population, including multiple sequence types and diverse OXA-51-like and ADC variants, further illustrates the complexity of *Acinetobacter* resistance epidemiology. Although NG-TEST^®^ *Acineto*-5^®^ cannot replace WGS for high-resolution epidemiological investigations, rapid detection of major carbapenemases could provide a useful frontline surveillance approach, with positive results prompting targeted molecular investigation when required.

An important strength was the assay’s performance beyond *A. baumannii*. Sensitivity and specificity reached 100% within the ACB complex, while sensitivity was 95.6% among non-ACB isolates, largely because one *A. radioresistens* carrying chromosomally encoded OXA-105 was not detected under the manufacturer’s recommended conditions. Four of five *A. radioresistens* isolates were detected at the recommended inoculum, whereas increasing the inoculum enabled detection of all five. Their low imipenem MICs and absence of detectable imipenem-hydrolytic activity suggest limited expression of the chromosomal OXA-23 variants (43). This finding indicates that assay performance depends not only on the genetic presence of a resistance determinant but also on its expression and the amount of bacterial material tested.

Another relevant finding was the broader recognition of certain OXA variants. OXA-143 and OXA-542 were detected through the OXA-24-targeted component, likely reflecting the sequence conservation with OXA-24. Although this extends the practical coverage of the assay, closely related OXA variants cannot necessarily be distinguished by LFIA; sequencing therefore remains necessary for precise epidemiological characterization.

Several limitations should be considered. First, only a subset of known variants was represented, limiting extrapolation to all currently described carbapenemases (Figure 4A-D). Nevertheless, the tested OXA variants encompassed broad phylogenetic diversity and showed >96% amino acid identity with their respective reference enzymes, while the NDM- and VIM-specific antibodies are identical to those used in the extensively evaluated NG-TEST^®^ CARBA-5^®^ assay (21, 33, 44) and have shown broad variant detection. Larger and geographically diverse collections will nevertheless be required to define the full breadth of variant coverage and analytical performance. Second, OXA-235-like and OXA-679-like, recently described acquired CHDLs, are not targeted by the current assay (37, 38). WGS therefore remains important when an LFIA result is negative despite a resistant phenotype or when an uncommon carbapenemase is suspected. Third, the study primarily evaluated cultured isolates and therefore does not establish assay performance directly on clinical specimens. Finally, although the assay demonstrated excellent analytical and biological performance, the study did not assess effects on patient outcomes, antimicrobial consumption, time to effective therapy, or infection-control interventions. Prospective clinical studies are needed to determine whether faster carbapenemase detection translates into measurable clinical, stewardship, or infection-control benefits.

In conclusion, NG-TEST^®^ *Acineto*-5^®^ demonstrated high concordance with WGS and reliable detection of clinically important carbapenemases in *Acinetobacter* spp. Its high sensitivity, specificity, broad species coverage, and ability to detect multiple carbapenemases provide potential value for clinical microbiology, antimicrobial stewardship, infection prevention, and surveillance. The assay should therefore be viewed as a rapid complementary method rather than a replacement for antimicrobial susceptibility testing or WGS, bridging the interval between isolation and definitive characterization of resistance.

## Data Availability

All data produced in the present work are contained in the manuscript

## Acknowledgments

S.O, H.V., K.J. and T.N. are members of the ESCMID Study Group for Antimicrobial Resistance Surveillance (ESGARS).

## Ethical approval

See section in Materials and Methods section

## Author Contributions

Development of the assay, A.C., L.F., M.L.Q., and C.M.; Conceptualization, K.J., S.S., H.V. and T.N.; methodology, K.J., H.V. and T.N.; validation, S.O., K.J., and T.N.; formal analysis, A.P., and S.O.; investigation, S.O., E.D., A.V. and A.P.; data curation, K.J. and T.N.; writing— original draft preparation, T.N.; writing—review and editing, All; supervision, S.S., K.J. and T. N.; project administration, K.J. and T.N. All authors have read and agreed to the published version of the manuscript.

